# The potential role of IL-6 in monitoring severe case of coronavirus disease 2019

**DOI:** 10.1101/2020.03.01.20029769

**Authors:** Tao Liu, Jieying Zhang, Yuhui Yang, Hong Ma, Zhengyu Li, Jiaoyu Zhang, Ji Cheng, Xiaoyun Zhang, Yanxia Zhao, Zihan Xia, Liling Zhang, Gang Wu, Jianhua Yi

**Affiliations:** Cancer center, Union Hospital, Tongji Medical College, Huazhong University of Science and Technology, Wuhan 430022, China; Department of Endocrinology, Union Hospital, Tongji Medical College, Huazhong University of Science and Technology, Wuhan 430022,China; Department of Gastrointestinal Surgery, Union Hospital, Tongji Medical College, Huazhong University of Science and Technology, Wuhan 430022,China; Liver intensive care unit, Zhongshan Hospital, Fudan University, Shanghai 200032, China; Department of Infectious Diseases, Union Hospital, Tongji Medical College, Huazhong University of Science and Technology, Wuhan 430022, China

**Keywords:** coronavirus disease 2019, interleukin-6

## Abstract

**Background:** The outbreak of coronavirus disease 2019 (COVID-19) in Wuhan City, China has spreads rapidly since December, 2019. Most patients show mild symptoms, but some of them develop into severe disease. There is currently no specific medication. The purpose of this study is to explore changes of markers in peripheral blood of severe COVID-19 patients, which may be of value in disease monitoring.

**Methods:** Clinical data of patients with nonsevere and severe type COVID-19 diagnosed by laboratory test in our institution were collected. The relationship between peripheral blood cells and cytokines, clinical manifestation and outcome was analyzed.

**Results:** A total of 69 severe type COVID-19 patients were included. On admission, the median age of severe cases was 56-year old, with 52.17% female patient. The most common symptoms were fever (79.72%), cough (63.77%), shortness of breath (57.97%) and fatigue (50.72%). Diarrhea is less common. The most common comorbidity is hypertension. Upon admission, the proportion of bilateral pulmonary involvement and interstitial abnormalities evidenced by chest computed tomography (CT) imaging in severe cases was 60.87% and 27.54%, respectively. Compared with patients with nonsevere disease, those with severe disease showed obvious lymphocytopenia. Elevated level of lactate dehydrogenase (LDH), C-reactive protein (CRP), ferritin and D-dimer was found in most cases. Two patients (2.9%) needed transfer to the intensive care unit. Baseline immunological parameters and most of the inflammatory parameters were basically within the normal range. However, baseline interleukin-6 (IL-6) was significantly increased in severe type, which was closely related to the maximal body temperature during hospitalization and to CT findings. Baseline IL-6 was also significantly related to the increase of baseline level of CRP, LDH, ferritin and D-dimer. The increase of baseline IL-6 level suggests that it may positively correlate with the severity of COVID-19. Among the 30 severe type patients whose IL-6 was assessed before and after treatment, significant decrease in IL-6 and improved CT assessment was found in 25 patients after treatment. Whereas the IL-6 level was further increased in 3 cases, which was closely related to disease progression. It is suggested that IL-6 may be used as a marker for disease monitoring in severe COVID-19 patients.

**Conclusions:** On admission, the baseline level of IL-6, CRP, LDH and ferritin was closely related to the severity of COVID-19, and the elevated IL-6 was significantly related to the clinical manifestation of severe type patients. The decrease of IL-6 was closely related to treatment effectiveness, while the increase of IL-6 indicated disease exacerbation. Collectively, the dynamic change of IL-6 level can be used as a marker for disease monitoring in patients with severe COVID-19.

## INTRODUCTION

In early December, 2019, the first cases of pneumonia of unknown origin were reported in Wuhan City, Hubei Province Province, China. The pathogen has been identified as a novel β-coronavirus by full-genome sequencing and is named severe acute respiratory syndrome coronavirus 2 (SARS-CoV-2) which shares phylogenetic similarity with severe acute respiratory syndrome coronavirus (SARS-CoV) that caused the outbreak of SARS in 2003.^1-3^ Disease caused by SARS-CoV-2, which has been highly contagious and spread rapidly nationwide and worldwide, has been designated coronavirus disease 2019 (COVID-19) by World Health Organization (WHO). Analysis of the epidemiological pattern curve of COVID-19 showed that the overall epidemic pattern was aggregation outbreak. Most cases are mild to moderate and curable, and the overall crude mortality rate is low.^2^ However, a proportion of patients with severe disease characterized by respiratory dysfunction have shown a high mortality rate.^2 4^ By February 16, 2020, a total number of 57934 cases have been reported in mainland China, including 10644 severe cases. As the outbreak area of COVID-19, the total number of confirmed cases in Hubei is 49847, with 9797 (19.6%) severe cases and an overall mortality rate of 3.4%. The disease incidence and mortality rate of severe COVID-19 in Hubei is relatively high as compared with other areas in China.

At present, due to the lack of reliable marker and effective antiviral medication, the monitoring of severe cases of COVID-19 mainly relies on the observation of clinical presentation.^4 5^ Previous studies have suggested that lymphocytopenia and inflammatory cytokine storm are typical abnormalities in infections caused by highly pathogenic coronavirus, such as SARS and MERS, and are considered disease severity related.^6-9^ Similarly, a decrease in lymphocyte count and an increase in inflammatory cytokines in peripheral blood have been reported in COVID-19 patients.^10-12^ Given the rapid spread of COVID-19 and the high mortality rate of severe cases, a better understanding of the clinical features is urgently needed and may help screen out reliable markers for inflammation monitoring through the course of disease. According to previous relevant literature on viral pneumonia and the current therapeutic experience on severe type COVID-19, the storm of inflammatory factors may be the main reason for rapid disease progression and poor treatment response.^6 7 13^ In this study, by collecting data of severe cases of laboratory-confirmed COVID-19 cases, we analyzed the clinical characteristics and inflammatory markers in patients with severe type COVID-19 in Wuhan City to explore potential markers for disease monitoring.

## METHODS

### Data source and collection

COVID-19 was diagnosed in accordance with the WHO interim guidance. A confirmed case of was defined as positive for SARS-CoV-2 nucleic acid on high-throughput sequencing or real-time reverse transcriptase polymerase chain reaction (RT-PCR) assays of nasal and pharyngeal swab specimens. Only laboratory-confirmed cases were included in this study, while disease diagnosed based on clinical presentation and imaging findings, but not on SARS-CoV-2 detection, were excluded. The severity of COVID-19 was classified according to the Guidelines for the Diagnosis and Treatment of Novel Coronavirus (2019-nCoV) Infection (Trial Version 5) issued by the National Health Commission of the People’s Republic of China. Severe case was defined when any of the following criteria was met: 1. dyspnea, respiration rate (RR) ≥30 times / min; 2. oxygen saturation by pulse oximeter ≤93% in resting state; 3. partial pressure of arterial oxygen (PaO_2_) to fraction of inspired oxgen (FiO_2_) ratio ≤300 mm Hg (l mm Hg=0.133kPa). We collected data of 69 patients with severe type COVID-19 hospitalized in the Department of Infectious Diseases, Union Hospital, Tongji Medical College, Huazhong University of Science and Technology, between January 21 and February 16, 2020. A retrospective study on the clinical characteristics and laboratory examination was conducted. 11 nonsevere COVID-19 patients were included for comparison. This study was approved by the Research Ethics Committee of Tongji Medical College, Huazhong University of Science and Technology. Verbal consent was obtained from patients before the enrollment. Written informed consent was waived due to the urgent need data collection.

The clinical symptoms, physical signs and results of laboratory examination of patients were recorded. Radiologic evaluation included chest computed tomography (CT) scan. Laboratory tests included baseline whole blood cell count, blood chemistry, coagulation test, C-reactive protein (CRP), procalcitonin (PCT), lactate dehydrogenase (LDH), ferritin, erythrocyte sedimentation rate (ESR), creatine kinase (CK), and lymphocyte subset and cytokine profile analysis on admission. In addition, cytokine profile follow-up was conducted in 30 severe type patients after treatment. Treatment plan are recorded. Time from the symptom onset to initial treatment, initial COVID-19 diagnosis, development of pneumonia evidenced by CT scan, and from the diagnosis of pneumonia to discharge were recorded.

### Laboratory diagnosis

Nasal and pharyngeal swab specimens were collected and placed into a collection tube containing preservation solution for virus^10^. Real-time RT-PCR assay for SARS-CoV-2 was conducted by the viral nucleic acid detection kit according to the manufacturer’s protocol (Shanghai bio-germ Medical Technology, Co. Ltd.). Laboratory confirmation of COVID-19 was performed by local Center for Disease Control and Prevention (CDC) in accordance with the Chinese CDC protocol. Whenever needed, specimens, including sputum or alveolar lavatory fluid, blood, urine and feces, were cultured to assess potential bacterial and/or fungal infection that might accompany with SARS-CoV-2 infection.

### Flow cytometry and ELISA detection

The lymphocyte test kit (Beckman Coulter Inc., FL, USA) was used for lymphocyte subset analysis by flow cytometry. Plasma cytokines (IL-2, IL-4, IL-6, IL-10, tumour necrosis factor (TNF)-α and interferon (IFN)-γ) were detected by ELISA with human Th1/2 cytokine kit II (BD Ltd., Franklin Lakes, NJ, USA). All tests were performed according to the product manual.

### Statistics

Continuous variables were described as means and standard deviations, or medians and interquartile range (IQR) values. Categorical variables were expressed as counts and percentages. Continuous variables were compared by the Mann Whitney U test. Proportions for categorical variables were compared by the *chi*-square test and Fisher’s exact test as appropriate. Correlations were determined by Spearman rank correlation analysis and Kendall correlation analysis. All statistical analyses were performed by Graphpad Prism (version 5.0) and SPSS 26.0 (IBM SPSS Statistics 26.0). For all statistical analysis, P<0.05 was considered statistically significant.

## RESULTS

### Demographic and clinical characteristics

A total of 69 severe cases were included. The median age was 56 years. Female accounted for 52.17% of the enrolled cases. The smoking rate was 11.59%. Fever (79.72%), cough (63.77%), shortness of breath (57.97%) and fatigue (50.72%) were the most common symptoms, while diarrhea (15.94%), vomiting (7.25%) and sore throat (5.80%) were relatively less common. 36.23% of patients had at least one comorbidity (e.g., hypertension, chronic obstructive pulmonary disease, coronary heart disease and hepatitis B virus infection), and there were four cases of tumor. Another 11 nonsevere cases were also enrolled (table 1).

**Table 1.** Clinical characteristics of patients with severe COVID-19

| Characteristics | All patients<br>(N = 80) | Nonsevere<br>(N = 11) | Severe<br>(N = 69) | P value |
| --- | --- | --- | --- | --- |
| Age - Median year (range) | 53.00<br>(26.00-86.00) | 31.00<br>(26.00-58.00) | 56.00<br>(27.00-86.00) | < 0.001 |
| Gender - No. (%) |  |  |  | < 0.037 |
| Male | 34 (42.50) | 1 (9.09) | 33 (47.83) |  |
| Female | 46 (57.50) | 10 (90.91) | 36 (52.17) |  |
| Smoking history - No. (%) |  |  |  | 0.591 |
| Non-smoker | 72 (90.00) | 11 (100.00) | 61 (88.41) |  |
| Smoker | 8 (10.00) | 0 (0.00) | 8 (11.59) |  |
| Fever on admission - No. (%) | 64 (80.00) | 9 (81.82) | 55 (79.72) | 0.808 |
| Temperature - No. (%) |  |  |  |  |
| < 37.5 □ | 17 (21.25) | 2 (18.18) | 15 (21.74) |  |
| 37.5-38.0 □ | 19 (23.75) | 4 (36.36) | 15 (21.74) |  |
| 38.1-39.0 □ | 38 (47.50) | 5 (45.45) | 33 (47.83) |  |
| > 39.0 □ | 6 (7.50) | 0 (0.00) | 6 (8.70) |  |
| Fever during hospitalization<br>- No. (%) | 46 (57.50) | 2 (18.18) | 44 (63.77) | 0.012 |
| Highest temperature during<br>hospitalization - No. (%) |  |  |  |  |
| < 37.5 □ | 35 (43.75) | 10 (90.91) | 25 (36.23) |  |
| 37.5-38.0 □ | 16 (20.00) | 1 (9.09) | 15 (21.74) |  |
| 38.1-39.0 □ | 23 (28.75) | 0 (0.00) | 23 (33.33) |  |
| > 39.0 □ | 6 (7.50) | 0 (0.00) | 6 (8.70) |  |
| Symptoms - No. (%) |  |  |  |  |
| Conjunctival congestion | 1 (1.25) | 0 (0.00) | 1 (1.45) | 1.000 |
| Nasal congestion | 5 (6.25) | 0 (0.00) | 5 (7.25) | 1.000 |
| Headache | 8 (10.00) | 0 (0.00) | 8 (11.59) | 0.516 |
| Cough | 53 (66.25) | 9 (81.82) | 44 (63.77) | 0.405 |
| Sore throat | 8 (10.00) | 4 (36.36) | 4 (5.80) | 0.009 |
| Sputum production | 15 (18.75) | 1 (9.09) | 14 (20.29) | 0.640 |
| Fatigue | 36 (45.00) | 1 (9.09) | 35 (50.72) | 0.024 |
| Hemoptysis | 2 (2.50) | 2 (18.18) | 0 (0.00) | 0.017 |
| Shortness of breath | 44 (55.00) | 4 (36.36) | 40 (57.97) | 0.312 |
| Nausea or vomiting | 7 (8.75) | 2 (18.18) | 5 (7.25) | 0.245 |
| Diarrhea | 15 (18.75) | 4 (36.36) | 11 (15.94) | 0.232 |
| Myalgia or arthralgia | 12 (15.00) | 0 (0.00) | 12 (17.39) | 0.296 |
| Shivering | 13 (16.25) | 0 (0.00) | 13 (18.84) | 0.257 |
| Physical signs - No. (%) |  |  |  |  |
| Throat congestion | 3 (3.75) | 0 (0.00) | 3 (4.35) | 1.000 |
| Tonsil swelling | 0 (0.00) | 0 (0.00) | 0 (0.00) |  |
| Lymph node enlargement | 0 (0.00) | 0 (0.00) | 0 (0.00) |  |
| Skin rash | 0 (0.00) | 0 (0.00) | 0 (0.00) |  |
| Coexisting disorders - No. (%) |  |  |  |  |
| Any of the following | 28 (35.00) | 3 (27.27) | 25 (36.23) | 0.812 |
| Diabetes | 11 (13.75) | 0 (0.00) | 11 (15.94) | 0.340 |
| Hypertension | 14 (17.50) | 0 (0.00) | 14 (20.29) | 0.223 |
| Coronary heart disease | 6 (7.50) | 0 (0.00) | 6 (8.70) | 0.589 |
| Cerebrovascular diseases | 0 (0.00) | 0 (0.00) | 0 (0.00) |  |
| Hepatitis B virus infection | 1 (1.25) | 0 (0.00) | 1 (1.45) | 1.000 |
| Cancer† | 7 (8.75) | 3 (27.27) | 4 (5.80) | 0.051 |
| Chronic renal diseases | 0 (0.00) | 0 (0.00) | 0 (0.00) |  |
| Immunodeficiency | 0 (0.00) | 0 (0.00) | 0 (0.00) |  |

† Cancer referred to any type of malignancy. All cases were stable disease.
P values denoted the comparison between nonsevere and severe cases.

### Radiologic and laboratory findings

Table 2 showed the radiologic and laboratory findings on admission. The most common CT patterns in severe cases upon admission were bilateral patchy shadowing (60.87%) and interstitial abnormalities (27.54%), while the common type mainly manifested as focal ground glass opacity and patchy shadowing (54.5%) (table 2).

**Table 2.**
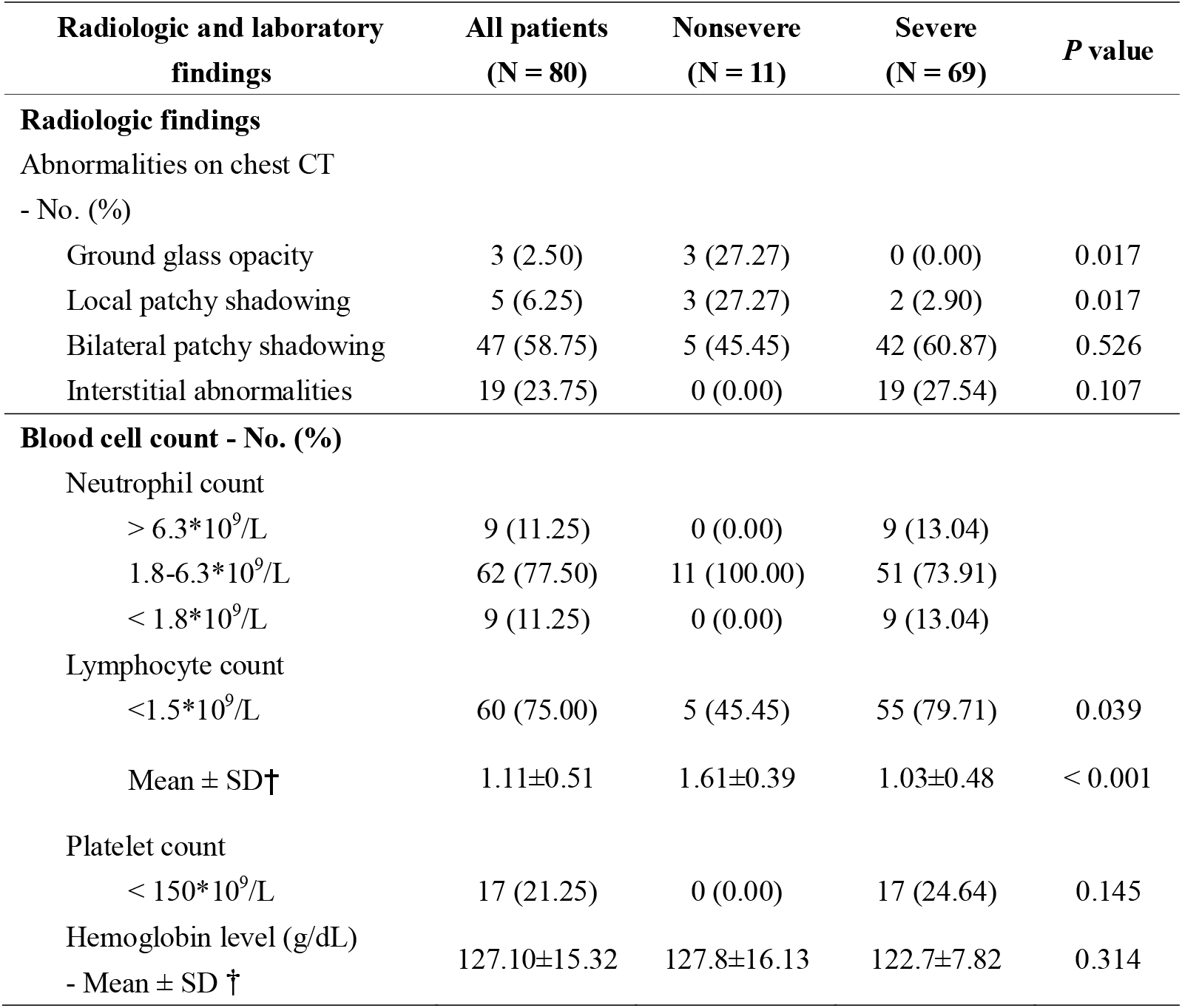
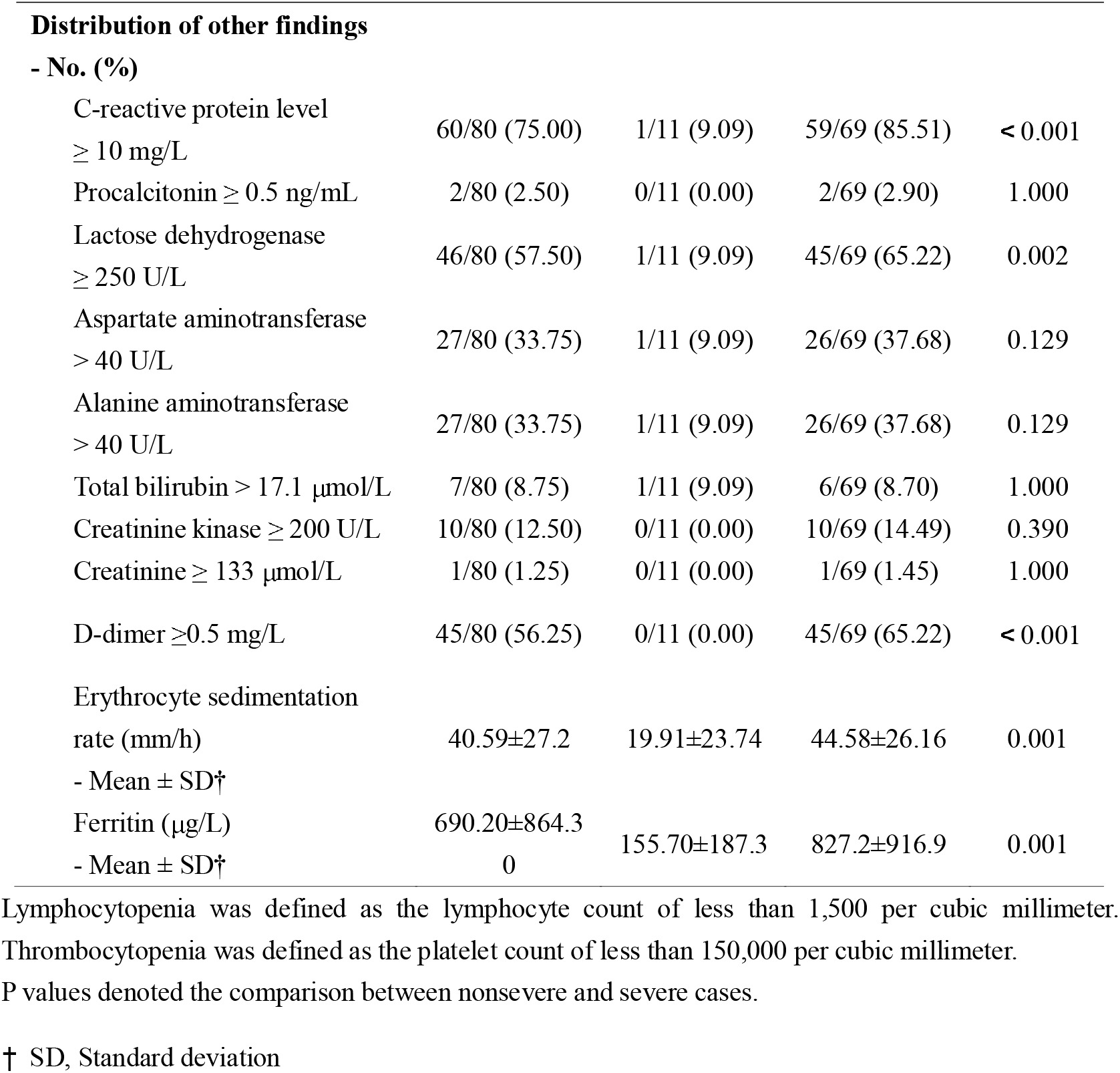
Radiologic and laboratory findings of patients with severe COVID-19

Baseline neutrocytopenia, lymphocytopenia and thrombocytopenia were observed in 13.04%, 79.71% and 24.64% of the severe type patients, respectively. Lymphocyte count in severe case was significantly less than the nonsevere cases (figure 1A). Increased creatine kinase was present in 14.49% of the cases. Elevated level of alanine aminotransferase and aspartate aminotransferase was more common and both was detected in 37.68% of the cases (table 2).

**Figure 1.**
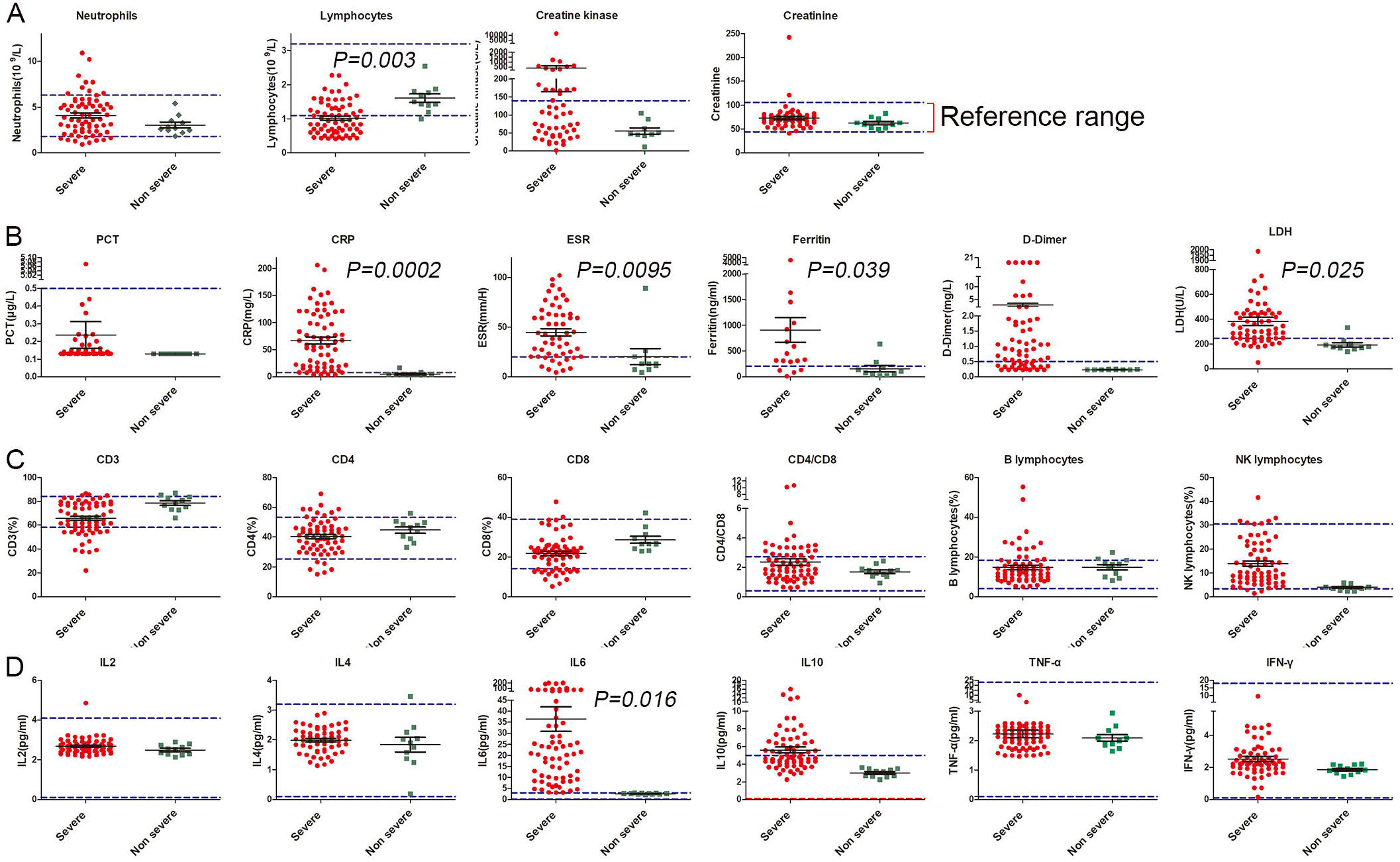
(A) Neutrophils and creatinine level from severe type COVID-19 patients were in the normal range. Compared with nonsevere cases, lymphocytes decreased while creatine kinase (CK) increased significantly in patients with severe COVID-19. (B) The procalcitonin (PCT) of patients with severe COVID-19 was basically normal, while the level of erythrocyte sedimentation rate (ESR), ferritin, C-reactive protein (CRP), D-Dimer and lactate dehydrogenase (LDH) was significantly increased. (C) Lymphocyte subgroup analysis showed that the proportion of CD4^+^ T cells, CD8^+^ T cells, B cells, natural kill(NK)cells and CD4^+^ T cells/CD8^+^ T cells ratio were within the normal range, and there was no significant difference between the two groups. (D) Cytokine profile analysis showed that compared with nonsevere type, there was no difference in the level of IL-2, IL-4, TNF-α and IFN-γ, while IL-10 increased slightly and IL-6 increased significantly in patients with severe COVID-19.

For inflammatory parameters, abnormal baseline PCT was not common in both nonsevere and severe patients. However, patients with severe disease showed more common and more prominent level of D-dimer, ESR, LDH, CRP and ferritin than those with nonsevere disease (figure 1B).

### Treatment and clinical outcomes

89.86% of the severe type patients received antibiotics, 63.77% received antiviral therapy (20.29% with oseltamivir, 52.17% with arbidol and 7.25% with lopinavir/ritonavir), 42.03% received glucocorticoids and 50.72% received human immunoglobulin (table 3). 55.07% of the severe cases needed oxygen therapy, among which 27.54% received high-flow oxygen therapy, non-invasive ventilation or invasive ventilation (table 3).

**Table 3.** Complications, treatment and clinical outcomes of patients with severe COVID-19

| **Characteristics** | **All patients**<br>**(N = 80)** | **Nonsevere**<br>**(N = 11)** | **Severe**<br>**(N = 69)** | **P value** |
| --- | --- | --- | --- | --- |
| **Complications - No. (%)** |  |  |  |  |
| Septic shock | 1 (1.25) | 0 (0.00) | 1 (1.45) |  |
| Acute respiratory distress<br>syndrome | 7 (8.75) | 0 (0.00) | 7 (10.14) |  |
| Acute kidney injury | 0 (0.00) | 0 (0.00) | 0 (0.00) |  |
| Disseminated intravascular<br>coagulation | 0 (0.00) | 0 (0.00) | 0 (0.00) |  |
| Rhabdomyolysis | 0 (0.00) | 0 (0.00) | 0 (0.00) |  |
| **Time to event - days** |  |  |  |  |
| From symptom onset to initial<br>treatment |  |  |  |  |
| Median | 1.00 | 1.00 | 1.00 |  |
| (interquartile range) | (1.00-4.00) | (1.00-5.00) | (1.00-4.00) |  |
| Mean ± SD† | 2.94±3.67 | 3.55±4.72 | 2.83±3.48 | 0.822 |
| From symptom onset to initial COVID-19 diagnosis |  |  |  |  |
| Median | 4.00 | 7.00 | 4.00 |  |
| (interquartile range) | (2.00-7.00) | (2.00-13.00) | (1.75-7.00) |  |
| Mean ± SD† | 5.23±4.44 | 7.36±5.52 | 4.83±4.14 | 0.141 |
| From symptom onset to development of pneumonia |  |  |  |  |
| Median | 4.00 | 8.00 | 4.00 |  |
| (interquartile range) | (2.00-7.50) | (2.00-13.00) | (2.00-7.00) |  |
| Mean ± SD† | 5.35±4.46 | 7.73±5.62 | 4.90±4.11 | 0.115 |
| From development pneumonia to recovery |  |  |  |  |
| Median | 18.00 | 18.00 | 18.00 |  |
| (interquartile range) | (16.00-23.00) | (12.25-22.50) | (16.00-23.00) |  |
| Mean ± SD† | 18.95±5.49 | 18.10±7.48 | 19.23±4.80 | 0.615 |
| **Treatments - No. (%)** |  |  |  |  |
| Antibiotics | 73 (91.25) | 11 (100.00) | 62 (89.86) | 0.585 |
| Oseltamivir | 20 (25.00) | 6 (54.55) | 14 (20.29) | 0.039 |
| Ribavirin, ganciclovir or peramivir | 47 (58.75) | 3 (27.27) | 44 (63.77) | 0.044 |
| Arbidol | 49 (61.25) | 10 (90.91) | 39 (52.17) | 0.066 |
| Antifungal medications | 10 (12.50) | 0 (0.00) | 10 (14.49) | 0.390 |
| Systemic glucocorticoids | 29 (36.25) | 0 (0.00) | 29 (42.03) | 0.019 |
| Nebulized interferon-α inhalation | 70 (87.50) | 10 (90.91) | 60 (86.96) | 0.902 |
| Lopinavir/ritonavir | 5 (6.25) | 0 (0.00) | 5 (7.25) | 1.000 |
| Oxygen therapy | 39 (48.75) | 1 (9.09) | 38 (55.07) | 0.007 |
| High-flow nasal cannula | 11 (13.75) | 0 (0.00) | 11 (15.94) | 0.340 |
| Mechanical ventilation |  |  |  |  |
| Invasive | 2 (2.50) | 0 (0.00) | 2 (2.90) | 1.000 |
| Non-invasive | 6 (7.50) | 0 (0.00) | 6 (8.70) | 0.589 |
| Use of extracorporeal membrane oxygenation | 0 (0.00) | 0 (0.00) | 0 (0.00) |  |
| Use of continuous renal replacement therapy | 0 (0.00) | 0 (0.00) | 0 (0.00) |  |
| Use of intravenous immunoglobulin | 36 (45.00) | 1 (9.09) | 35 (50.72) | 0.024 |
| **Clinical outcomes** |  |  |  |  |
| Intensive care unit admission | 3 (3.75) | 0 (0.00) | 3 (4.35) | 1.000 |
| Death | 0 (0.00) | 0 (0.00) | 0 (0.00) |  |
| Recovery | 47 (58.75) | 10 (90.91) | 37 (53.62) | 0.022 |
| Hospitalization | 33 (41.25) | 1 (9.09) | 32 (46.38) | 0.022 |

† SD, Standard deviation

Among the enrolled patients with severe disease, 53.62% were cured and discharged, 46.38% were still hospitalized, three patients (4.35%) needed transfer to the intensive care unit and no death case occurred (table 3). Seven patients developed acute respiratory distress syndrome (ARDS) and one got septic shock (table 3). Compared with patients with nonsevere disease, the time from symptom onset to initial COVID-19 diagnosis and to development of pneumonia in patients with severe disease was shorter, but it was not significant (P>0.05) (figure 2A, B). There was no significant difference in the time from symptom onset to treatment, and the time from development of pneumonia to recovery in patients with severe disease was longer (P >0.05) (figure 2C, D).

**Figure 2.**
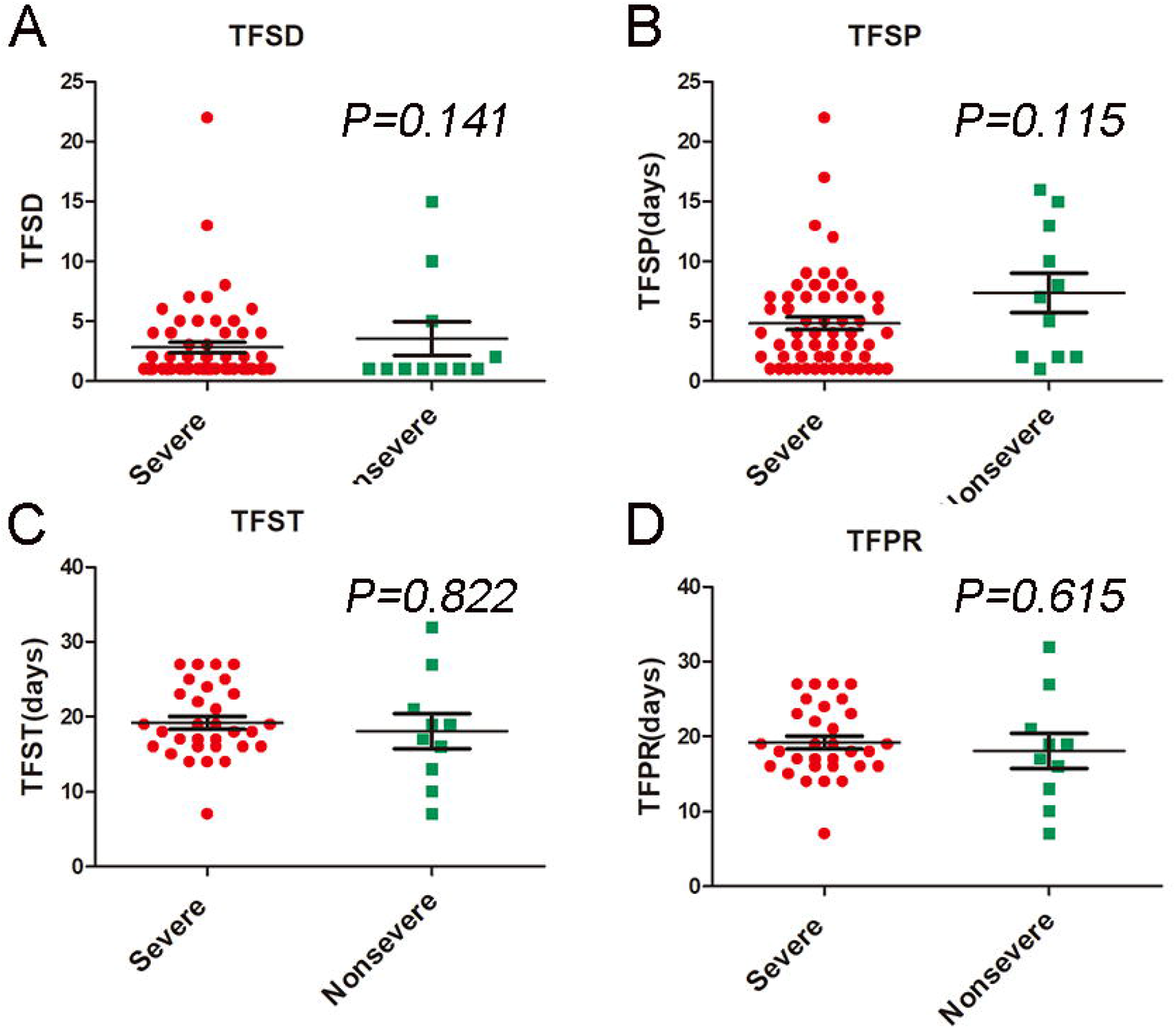
Compared with nonsevere COVID-19 patients, (A) the time from symptom onset to initial diagnosis (TFSD) and (B) the time from symptom onset to development of pneumonia (TFSP) of patients with severe COVID-19 was shorter, but there was no significant difference (P>0.05). There was no difference in (C) the time from symptom onset to treatment (TFST) and (D) the time from development of pneumonia to recovery (TFPR) of patients with severe COVID-19 (P>0.05).

### Immunological findings

The baseline proportion of CD4^+^ T cells, CD8^+^ T cells, B cells, natural killer (NK) cells, and the CD4^+^ T cells/CD8^+^ T cells ratio are basically within the normal range (figure 1C). The baseline level of IL-2, IL-4, IL-10, TNF-α and IFN-γ was within normal range, while IL-10 was slightly increased (figure 1D).

The IL-6 level was increased significantly on admission in severe cases as compared with the nonsevere cases (figure 1D). The baseline elevated IL-6 was positively correlated with the bilateral and interstitial pulmonary involvement (r=0.453, P=0.001) and closely related to the maximal body temperature during hospitalization (r=0.521, P=0.000) (figure 3A), Meanwhile, it was significantly related to the increase of CRP (r=0.781, P=0.001), LDH (r=0.749, P=0.001), ferritin (r=0.606, P=0.001) and D-dimer (r=0.679, P=0.001) (figure 3B-E). We found tendency showing that, on admission, the lower the IL-6 level, the shorter the time lapse from diagnosis to cure (r=0.049, P=0.763), whereas the higher the IL-6 level, the shorter the time lapse from symptom onset to pneumonia diagnosis (r= -0.116, P=0.345). The elevated level of IL-6 was associated with the administration of glucocorticords (r=0.301, P=0.001), human immunoglobulin (r=0.147, P=0.118), high flow oxygen inhalation (r=0.251, P=0.007), mechanical ventilation during hospital (r=0.223, P=0.017).

**Figure 3.**
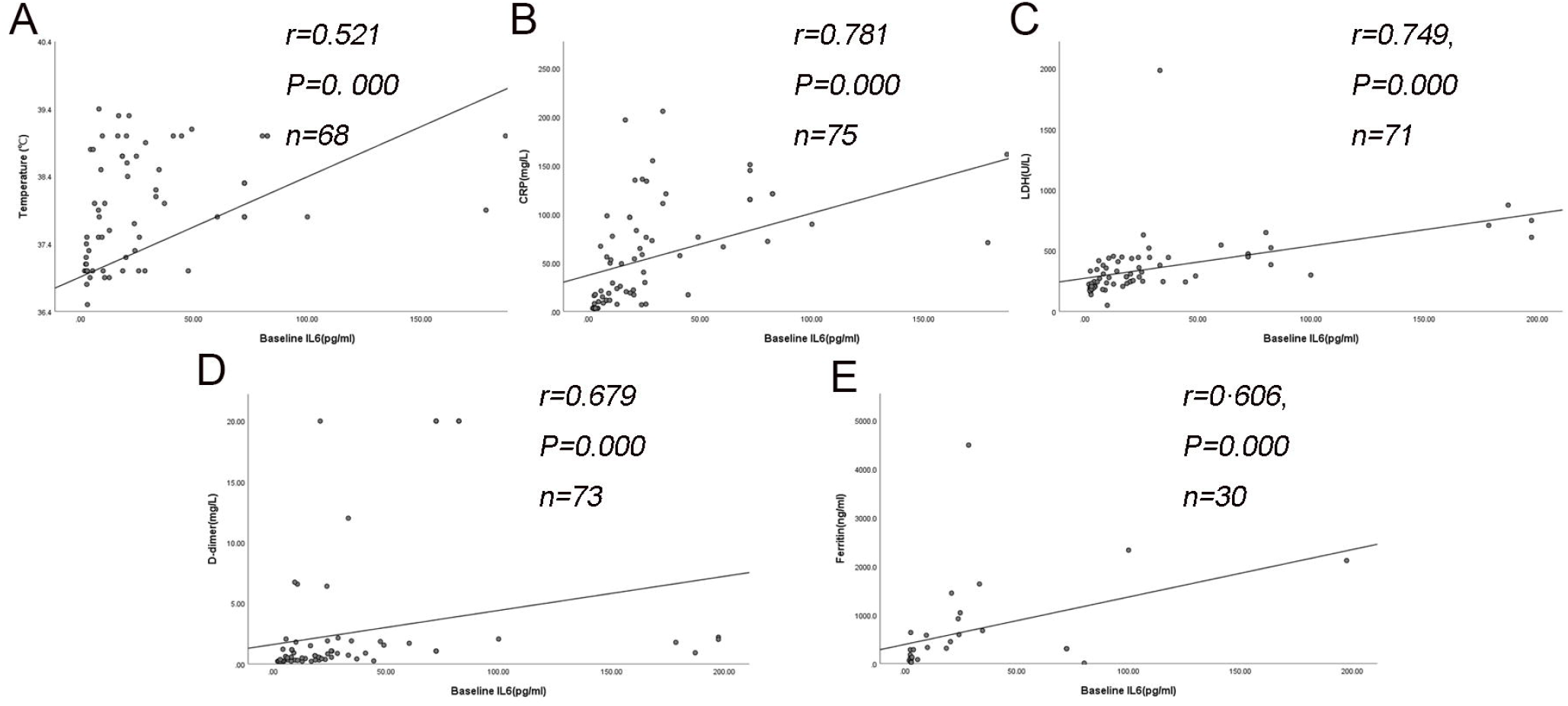
Correlation between baseline IL-6 level and clinical features of severe COVID-19 patients. (A) The baseline IL-6 level was positively correlated with the maximal body temperature during hospitalization. (B-E) The baseline IL-6 level was positively correlated with C-reactive protein (CRP), lactate dehydrogenase (LDH), ferritin and D-dimer.

Among the 30 patients whose IL-6 was assessed before and after treatment, 26 patients showed significantly reduced IL-6, which is accompanied with improved CT assessments after treatment (figure 4A). Disease of the other four patients exacerbated progressively. Among them, three patients showed persistent increase of IL-6 (figure 4B), while the change in IL-6 in one patient was not obvious. For this patient, there was an increase in PCT level, and the progression of lung disease was confirmed as bacterial, but not viral pneumonia, as evidenced by sputum culture positive for bacterial infection (figure 4C). Eventually, with the remission of disease (figure 5), the IL-6 level of this patient decreased significantly, and the D-dimer, CRP and LDH level also decreased significantly, all of which were associated with the resolution of clinical manifestation and CT images.

**Figure 4.**
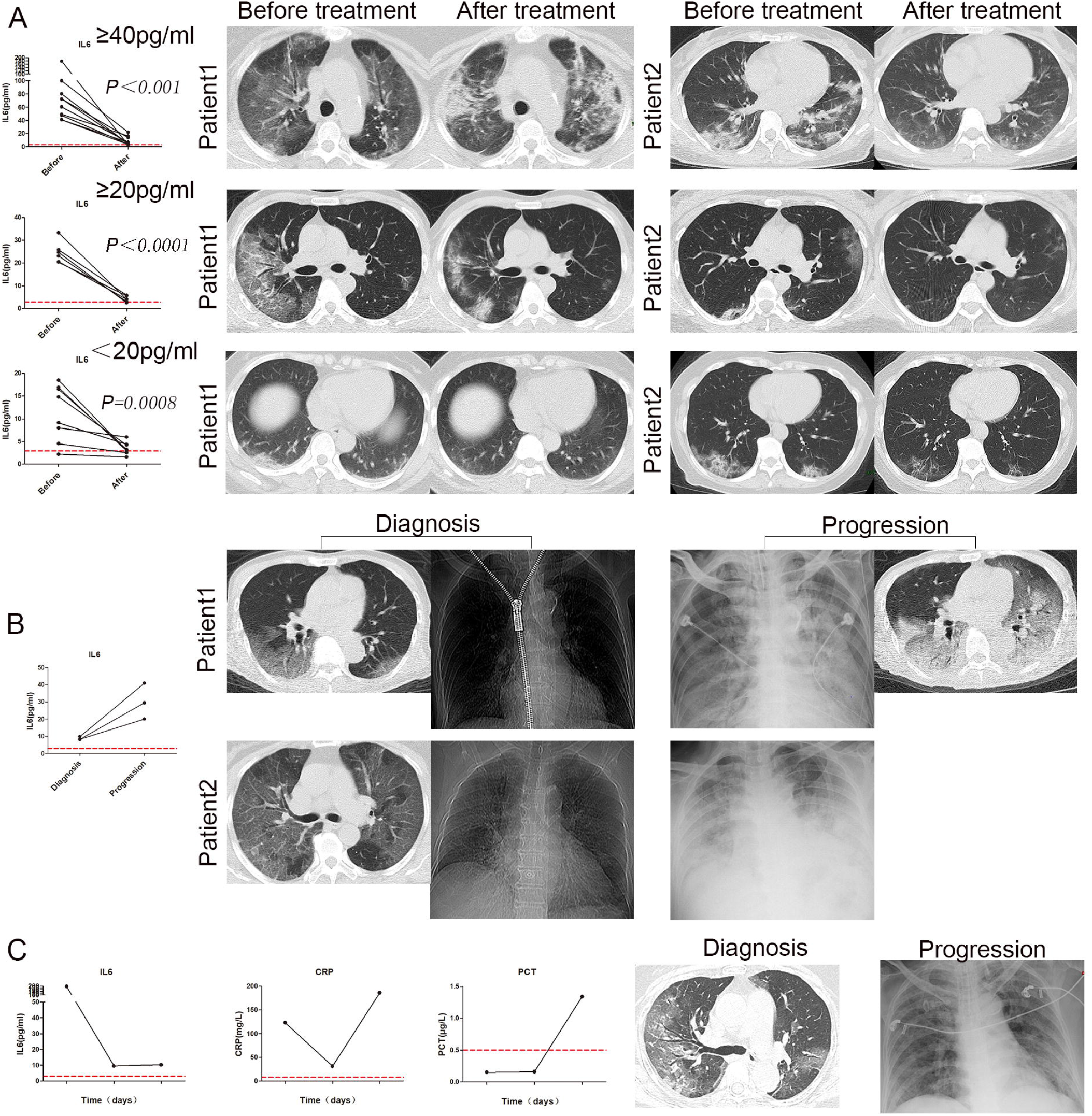
Variation in IL-6 and chest computed tomography in severe COVID-19 patients. (A) The elevated baseline IL-6 was correlated with the severity assessed by chest computed tomography (CT) scan, and the decrease in IL-6 after treatment was positively correlated with the improvement in chest CT images. (B) Three patients showed elevated IL-6 after treatment, which is associated with disease exacerbation and progressed CT imaging. (C) The baseline IL-6 was 197.39 pg/mL in a 69-year-old female patient who showed high fever and dyspnea. IL-6 decreased to 9.47 pg/mL after treatment, but the symptoms were not relieved. CT scan was not performed due to poor general condition, whereas chest X-ray showed aggravated pneumonia. The C-reactive protein (CRP) rebounded and procalcitonin (PCT) increased significantly. Follow up sputum culture confirmed the exacerbation was caused by bacterial infection.

**Figure 5.**
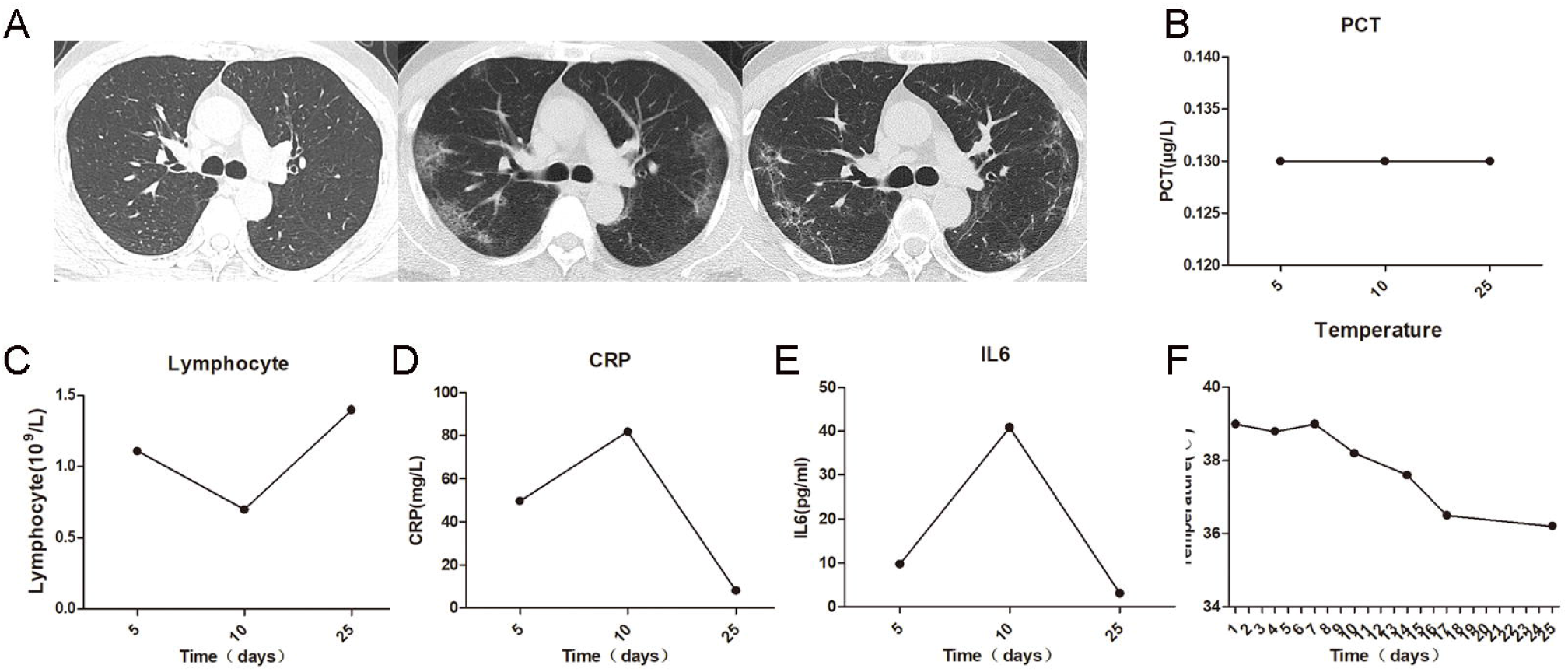
A 59-year-old male patient was diagnosed as COVID-19 on the fifth day from the onset of fever. (A) The chest computed tomography (CT) was still normal five days after symptom onset; and the patient presented with initial disease aggravation evidenced by CT scan ten days after symptom onset displaying bilateral multiple patchy ground glass opacities, and subsequent alleviation evidenced by improved CT images 25 days after symptom onset. (B) The procalcitonin (PCT) level had stayed in the normal range throughout the course of disease. (C) The lymphocyte count was still normal five days after symptom onset. It reached nadir and returned to normal during the disease course. (D-E) Similar trend of fluctuation was detected in the level of C-reactive protein (CRP) and interleukin-6 (IL-6). Both abnormal CRP and IL-6 were detected five days after symptom onset when the chest CT was still normal. In association with changes in CT scans, both CRP and IL-6 peaked ten days after symptom onset and returned to normal 25 days after symptom onset. (F) The body temperature correlated with the variation in CRP and IL-6, as it stayed abnormal during the rising phase while returned to normal during the decline phase of CRP and IL-6.

## DISCUSSION

This study shows that most of the severe cases of COVID-19 have the initial symptoms of fever, cough, shortness of breath and fatigue, while diarrhea is not common. Compared with the nonsevere cases, the imaging findings mainly involve bilateral and interstitial abnormalities. In patients with severe disease, more intensive and supportive treatment, including glucocorticoids, human immunoglobulin, interferon-α, antibiotics, antiviral therapy (oseltamivir and abidol) and oxygen therapy, was administered and relieved symptoms were observed in most cases. The time from initial symptoms to pneumonia in severe COVID-19 patients was shorter, the median recovery time was longer, and the onset of severe type was more rapid than the nonsevere patients.

Previous studies have suggested that lymphocytopenia and inflammatory cytokine storm are associated with the severity of infections caused by highly pathogenic coronavirus.^4 6 7 14 15^ Cytokines are signaling peptides, proteins, or glycoproteins that are secreted by many cell types, including immune, epithelial, endothelial, and smooth muscle cells. Cytokines allow context-dependent communication within the body.^13^ If the interactions that lead to cytokine production are destabilized, a “cytokine storm” can result, producing unbridled inflammation within tissues and key organs.^13^ Cytokine storms are associated with sepsis and septic shock, influenza, acute respiratory distress, and toxic response to medication and so on.^6 9 15^ IL-1, IL-6, IL-10, and TNF-a have been implicated in the 1918 Spanish flu pandemic, the 2003 SARS outbreak and the H5N1 avian influenza infections firstly recognized in 1987.^6 15 16^ Similarly, recent studies on COVID-19 patients have also reported a decrease in peripheral blood lymphocyte count and an increase in serum inflammatory cytokine.^10 11^ Cytokine storms, which can rapidly cause single or multiple organ failure and ultimately can be life-threatening, are considered to be an important cause of death in patients with severe COVID-19. SARS-CoV-2 infection can rapidly activate pathogenic T cells and produce granulocyte-macrophage colony stimulating factor (GM-CSF) and IL-6. GM-CSF will further activate CD14^+^CD16^+^ inflammatory monocytes and produce more IL-6 and other inflammatory factors, resulting in a cytokine storm that causes severe immune damage to the lungs and other organs.^17^

Results of this study indicated that compared with the common type, the changes in immunological parameters and cytokine level in severe type COVID-19 are inconspicuous. There was mild variation in IL-2, IL-4, IL-10, TNF-a, IFN-γ before and after treatment, all of which fluctuated within the normal range. However, elevated CRP, ferritin, IL-6 and LDH were associated with more intensive and prolonged treatment, which included glucocorticoids, human immunoglobulin, stronger antibiotics, high flow oxygen therapy or mechanical ventilation. Collectively, it showed that the above mentioned parameters were closely related to disease severity.

Further analysis showed that the changes in IL-6 were closely related to the disease. In this study, CRP, ferritin and IL-6, LDH decreased significantly after recovery. In association with disease progression evidenced by exacerbating pulmonary lesions on chest CT scan, IL-6 increased to a further degree, suggesting that IL-6 might be a valuable candidate for monitoring severe type COVID-19. To our interest, in one patient whose IL-6 level remained low as disease exacerbated, progression of pulmonary lesions was caused by bacterial infection, which might suggest the specificity of IL-6 in COVID-19.

IL-6 is synthesized by a variety of cells in the lung parenchyma, including alveolar macrophages, type II pneumocytes, T lymphocytes and lung fibroblasts. IL-6 is a pleiotropic cytokine important in regulating immunologic and inflammatory responses.^9 18^ IL-6 being an acute phase inflammatory cytokine suggests that measuring circulating IL-6 may reflect the inflammatory state of the lungs.^7 18^ This is supported by the often observed increase of IL-6 in ARDS acute complications of lung transplantation.^9 12 19 20^ Our findings that IL-6 elevated upon diagnosis and varied correspondingly accordance with disease outcomes support a shared mechanism of cytokine-mediated lung injury caused by virus. Siltuximab and tocilizumab are monoclonal antibodies (mAb) targeted against IL-6 and its receptor (IL6R).^21 22^ Both siltuximab and tocilizumab have been used to treat cytokine release syndrome following chimeric antigen receptor–armed T cells (CART-19) therapy for leukemia. Since we have found that IL-6 is related to COVID-19 severity, we suggest that targeting IL-6 may ameliorate cytokine storm-related symptoms in severe COVID-19 cases.^22 23^

In conclusion, severe COVID-19 patients require more intensive treatment, and the prognosis is relatively poor. IL-6 is positively correlated with disease severity, since it decreases with the remission while increases with the aggravation of the disease. Therefore, IL-6 may be an ideal marker of the disease monitoring. Targeting IL-6 may be effective in treating inflammatory cytokine storm during disease progression. A better understanding of the precise role of IL-6 in the pathogenesis of COVID-19, especially in the severe cases, may help us manage the disease.

## Data Availability

All data included in this study are available upon request by contact with the corresponding author.

## Acknowledgements

We thank the patients and their family for their enthusiastic participation in this study. We thank all the doctors, nurses and staff who are fighting campaign against COVID-19.

## Contributors

TL and JY conceptualized and designed the study, had full access to all data, and took responsibility for data integrity and accuracy of the analysis. JZ, YY, LZ, HM, ZYL wrote the manuscript. LZ, GW and JY reviewed the manuscript. TL, JZ and JC performed the statistical analysis. All authors contributed to data acquisition, analysis and interpretation, and approved the final version for submission.

## Patient consent for publication

Not required

## Ethics approval

The human study was approved the Research Ethics Committee of Tongji Medical College, Huazhong University of Science and Technology.

